# Interprofessional education curriculum and the knowledge of interdisciplinarity and multidisciplinary among undergraduate dental students

**DOI:** 10.64898/2026.08.13.26360104

**Authors:** Mario Brondani, Jonathan Rafael Garbim, Bruna Brondani, Vincent Lee, Abiola Adeniyi

**Affiliations:** Department of Oral Health Sciences, Faculty of Dentistry, University of British Columbia. 116/2199 Wesbook Mall. Vancouver, BC. V6T 1Z3. Canada; Department of Orthodontics and Pediatric Dentistry, School of Dentistry, University of Sao Paulo, São Paulo (SP), Brazil; Department of Pediatric Clinic (DCI) at the Ribeirão Preto School of Dentistry, University of São Paulo, São Paulo, Brazil; Department of Oral Health Sciences, Faculty of Dentistry, University of British Columbia. 370/2199 Wesbook Mall. Vancouver, BC. V6T 1Z3. Canada; School of Population and Global Affairs, Silberman College of Business, Fairleigh Dickinson University Vancouver. 247/842 Cambie St., Vancouver, BC V6B 2P6. Canada

**Keywords:** Interprofessional Education, Interdisciplinary, Multidisciplinary, Undergraduate dental education, Cross-sectional study

## Abstract

**Objectives:** Collaboration among health care professionals and the services they provide can be strengthen by interprofessional education (IPE). IPE can be implemented at the undergraduate level. Accordingly, the objective of the present study was to evaluate senior students’ understanding of the terms interdisciplinarity and multidisciplinarity within the context of IPE.

**Methods:** A retrospective cross-sectional study design was used. Students’ understanding of interdisciplinarity and multidisciplinarity was assessed through an assessment question completed by three consecutive cohorts of senior undergraduate dental students at the UBC Faculty of Dentistry between 2021–22 and 2023–24 (N = 177). Responses had a maximum of 100 words and were categorized into one of four predetermined themes: concordant knowledge (when both definitions were correct), discordant rhetoric (when both definitions were incorrect), switched ideas (when the definitions were reversed), and altered discourse (when the concepts of discipline and specialty were conflated). Descriptive and inferential statistical analyses were performed using SPSS Version 31®.

**Results:** Of the 177 students enrolled, 164 provided responses to the question on multidisciplinarity and interdisciplinarity: 60 students in 2021–22, 51 students in 2022– 23, and 53 students in 2023–24; the mean age was 25 years and 88 were female. Of the four predetermined themes, 45.7% of responses reflected concordant knowledge, 15.9% discordant rhetoric, 18.3% switched ideas, and 20.1% altered discourse. The logistic regression analysis showed age associated with a higher probability of providing the correct definitions (adjusted OR = 1.39; 95% CI: 1.06–1.83; p = 0.017).

**Conclusions:** Knowledge of interdisciplinary and multidisciplinary appeared to be retained by the majority of students. However, dental education programs, alongside other health professional training programs, should continue to incorporate interprofessional education through both didactic and experiential learning opportunities to equip students with the skills necessary to provide collaborative care in future practice.

## Introduction

Collaborative practice in health care can be fostered via interprofessional education (IPE), and remains fundamental to contemporary healthcare. It aims to improve patient outcomes, enhance safety in care delivery, and optimize the use of increasingly scarce resources.[1] IPE involves students from two or more health professional backgrounds learning about, from, and with one another, thereby promoting a clearer understanding of their respective roles and collaborative responsibilities.[2] It primarily refers to the integration of various individuals and professions and prepares team members to understand, respect, and communicate across their distinct roles.

Interdisciplinarity grounds IPE, referring to the integration of knowledge and methods whereby multiple health disciplines analyze, synthesize, and harmonize connections between their fields into a coordinated whole.[3] Interdisciplinarity challenges traditional silos of practice by fostering reciprocal interactions, in which professionals develop a combined, overlapping approach that generates new knowledge, perspectives, and shared problem-solving frameworks.[4] Multidisciplinarity, by contrast, refers to an independent, parallel approach in which different disciplines work on the same problem but do so separately, operating strictly within their own professional boundaries. In this model, information is gathered and analyzed in isolation, with professionals interacting ‘at-a-distance’ (e.g., through referrals, submission of separate reports, and so on) and with little to no overlap in their day-to-day activities.[3] Multidisciplinarity is not conducive to IPE, as effective collaboration must be built on mutual respect and clearly defined shared responsibilities, thereby preparing trainees for the real-world delivery of team-based care.[5]

Interprofessional collaborative practice centers on promoting the health and well-being of patients, including their oral health. Historically, however, oral health has largely been addressed in a siloed manner, effectively separated from the rest of the body. With the majority of oral health care practices working under a private system, care is often delivered through multidisciplinary approaches at best, mainly via referrals or brief consultations when needed.[6] Integrating oral healthcare providers into interprofessional teams enhances healthcare delivery and patient outcomes, given the well-established connections between oral and systemic health.[7],[8] However, significant barriers to this integration remain, including the training of oral health providers within rigid curricula deeply rooted in professional hierarchies,[9] limited faculty expertise in IPE,[10],[11] the historical evolution of dentistry as a profession distinct from medicine, and fragmented healthcare systems that hinder collaboration.[12],[13]

Nonetheless, IPE has been incorporated into some undergraduate dental training programs,[6,9] including in Canada.[10] For example, the University of British Columbia offers a two-year mandatory IPE curriculum for undergraduate dental students in Years 1 and 2 of the program (out of four years), delivered alongside students from 13 other disciplines, including dental hygiene, medicine, nursing, pharmacy, and occupational therapy.[14] This curriculum incorporates virtual learning environments, structured workshops, and a range of educational strategies, from classroom-based case discussions to immersive simulations and digital platforms that replicate clinical scenarios, thereby supporting a shift from multidisciplinary to interdisciplinary practice.[15] However, the knowledge and approaches gained within IPE curricula may dissipate if not re-assessed and reinforced throughout the curriculum. And may even disappear altogether if not applied in practice, particularly given the limited hands-on opportunities available for dental students to engage in interprofessional care during their training. Consequently, the conceptualization and application of inter- and multidisciplinarity may lost their relevance and fade-out over time.[16]

In this context, the objective of the present study was to evaluate senior students’ understanding of the concepts of interdisciplinarity and multidisciplinarity within IPE after completing such a curriculum in their junior years, before they become fully immersed in clinical care. This study contributes to ongoing efforts to advocate for sustained IPE and collaborative practice as essential components for enhancing communication, collaboration, and teamwork among oral health professionals and other healthcare providers.

## Methods

Ethical approval was received from the University of British Columbia (UBC) Behavioural Research Ethics Board (H25-01322). This retrospective cross-sectional design examined retained knowledge regarding the meaning and understanding of interdisciplinarity and multidisciplinarity in the provision of collaborative oral healthcare among three cohorts of senior undergraduate dental students at the UBC Faculty of Dentistry.

The IPE curriculum took place from September to December during Years 1 and 2 of the undergraduate dental program, designed to help students develop proficiencies required for team-based care. It includes a series of learning modules focused on complex health topics, enabling students to build collaborative skills, and spans more than 40 hours of instruction. This curriculum is mandatory for dental students and, although first launched in 2001 at university level, was formally implemented as an integrated curriculum in dentistry in 2015–2016 academic year under the evolving course *Principles of Ethical Practice*.[17,18]

In the spring of Year 3, approximately 36 months after their final IPE session, students revisit the concepts of interdisciplinarity and multidisciplinarity as they become fully immersed in hands-on clinical care in community clinics [19,20] and nursing homes.[21,22] At that time, they complete a 30-question assessment evaluating retained knowledge; questions include both multiple-choice and short-answer formats. The assessment is conducted in a computer lab using desktop computers with a lockdown browser on the Canvas® platform, without access to external materials. One question asks students to explain their understanding of these two concepts in 100 words or fewer: “*During your undergraduate training, interdisciplinary (or interdisciplinarity work) and multidisciplinary (or multidisciplinarity work) were concepts discussed. In the space below, explain your understanding of these two concepts in the provision of oral health care. You can use definitions, examples, or any other means to demonstrate your understanding. You may use up to 100 words*.” No other question in the assessment addressed these concepts. A maximum of two hours was allotted for completion of the entire assessment, graded on a scale from 0 to 10 and accounts for 25% of the course grade. Students may choose whether to answer all questions, with implications for their final grade.

The data consisted of responses to this specific question submitted by three consecutive cohorts of third-year dental students at UBC: 2021–2022 (63 students), 2022–2023 (55 students), and 2023–2024 (59 students), for a total of 177 participants. Responses were analyzed descriptively by gender, age, and cohort year. Each response was categorized into one of four pre-determined themes: (1) *concordant knowledge*, when responses reflected accurate understanding of both concepts; (2) *discordant rhetoric*, when responses did not reflect accurate understanding of one or both concepts; (3) *switched ideas*, when the understanding of both concepts were reversed; and (4) *altered discourse*, when responses referred to disciplines as the specialties within the same field, not as a distinct health profession.

As in our previous studies,[16,21,23] the analysis focused on categorizing textual data to interpret meanings and relationships among words and ideas within these above pre-determined themes.[24] Given the 100-word limit per response, all submitted answers were included in the analysis. Responses were securely downloaded from the Canvas® platform to the first author’s (MB) university computer, and three identical digital copies were created for analysis. Data were anonymized by removing student names and identification numbers; only academic year, age and gender were retained. Descriptive statistics (frequencies, percentages, and averages) were calculated using IBM SPSS Statistics (Version 31.0). Because all responses were included, the concept of data saturation was not applicable.

Three authors (MB, JRG, BB) independently analyzed an initial set of 15 responses to establish consensus. They subsequently met to compare their categorizations, while inter-rater reliability was assessed using Cohen’s kappa (κ) to quantify agreement beyond chance. Kappa value of 0.86 was obtained, referring to near-perfect agreement (95%CI 0.45-1.00). Following that level of agreement, the three authors independently analyzed separate subsets of the remaining digital data under the assumption that sufficient calibration had been achieved. The four themes are presented with selected student responses for illustrative rather than exhaustive purposes; responses were copied and pasted verbatim.

## Results

Of the 177 students enrolled, 164 responded to the question on multidisciplinarity and interdisciplinarity: 60 students in 2021–22, 51 in 2022–23, and 53 in 2023–24. Among these 164 respondents, 24 were female (45.3%) in 2023–24, 29 (56.9%) in 2022–23, and 35 (58.3%) in 2021–22, and the mean age was 25 ± 1.2 years. No statistically significant differences were observed in gender distribution or age across the three cohorts (Table 1).

**Table 1.** Characteristics of participants according to gender, age and responses per class year.

|  | **Class**<br>**2023-24** | **Class 2022-**<br>**23** | **Class 2021-**<br>**22** | **Total** | **p-value** |
| --- | --- | --- | --- | --- | --- |
| **Participants, n** | 53 | 51 | 60 | 164 |  |
| **Age (years), mean (SD)** | 25 (1.3) | 24 (1.1) | 25 (1.2) | 25 (1.2) | 0.071 |
| **Gender, n (%)** |  |  |  |  | 0.738 |
| Female | 24 (45.3) | 29 (56.9) | 35 (58.3) | 88 (53.7) |  |
| Male | 29 (54.7) | 22 (43.1) | 25 (41.7) | 76 (46.3) |  |
| **Themes (%)** |  |  |  |  | 0.189 |
| Concordant knowledge | 28 (52.8) | 25 (49.0) | 22 (36.7) | 75 (45.7) |  |
| Discordant rhetoric | 7 (13.2) | 7 (13.7) | 12 (20.0) | 26 (15.9) |  |
| Switched ideas | 8 (15.1) | 13 (25.5) | 9 (25.0) | 30 (18.3) |  |
| Altered discourse | 10 (18.9) | 6 (11.8) | 17 (28.3) | 33 (20.1) |  |

\*SD – Standard deviation. P-values were obtained using one-way ANOVA for age and Pearson's chi-square test for categorical variables.

**Table 2.**
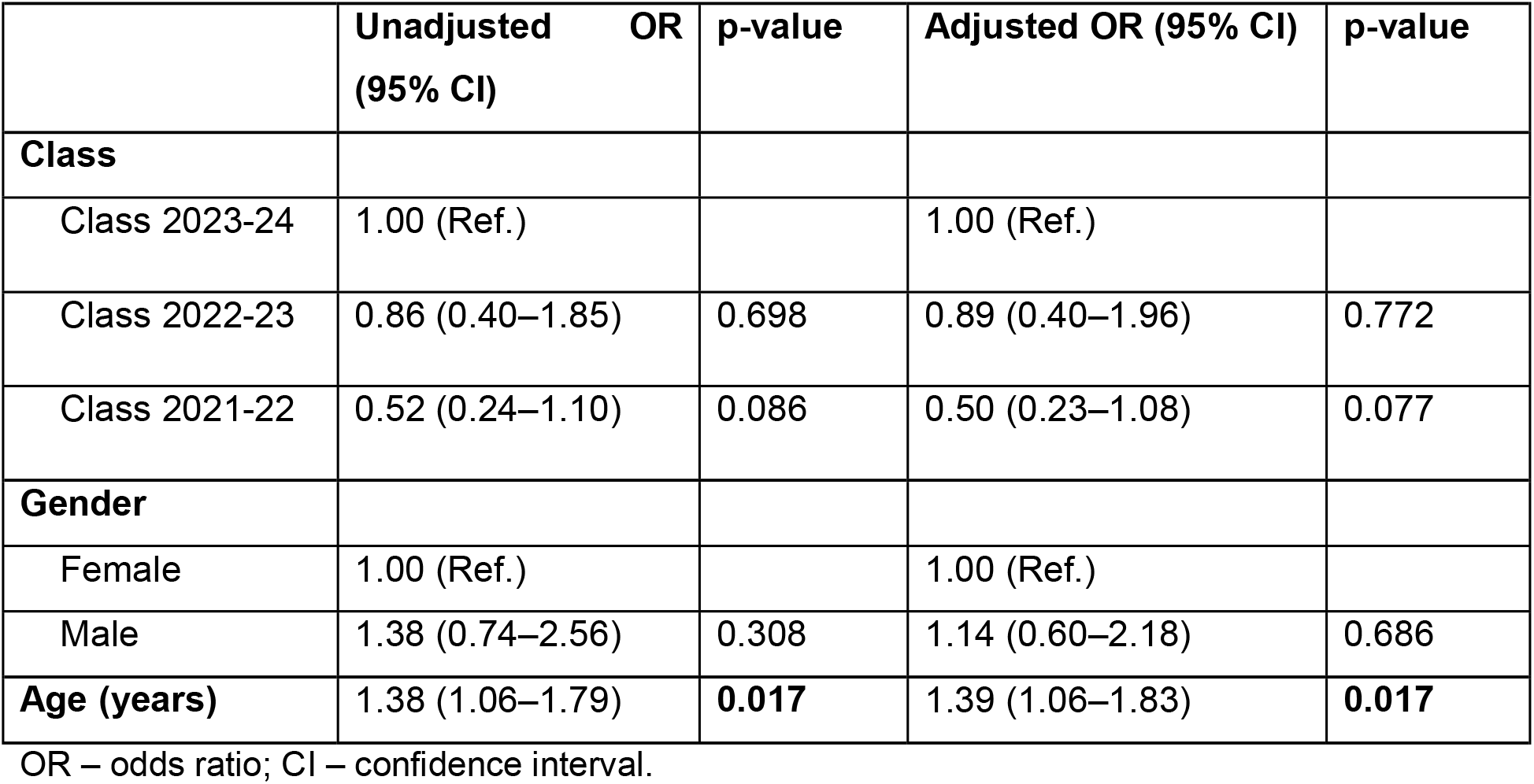
Unadjusted and adjusted logistic regression analyses of factors associated with providing the correct definition of interdisciplinary care (Theme 1).

Analysis of the 164 responses revealed concordant knowledge in 45.7% of time (36.7% in 2021–22, 49.0% in 2022–23, and 52.8% in 2023–24) (Table 1). In these cases, students correctly defined and/or exemplified both concepts, as illustrated below.

> “*Interdisciplinary emphasizes more on the interaction among different disciplines that are involved. this concept treats all participating disciplines as a dynamic entity. Multidisciplinary states the facts that more than one discipline is participating in the diagnosis and treatment of oral disease but it doesn’t imply their interactions. this is a more parallel concept*” (Male student, 2023-24).
>
> *“Interdisciplinary: when team members work together with the same goal. team members can have meetings with each other where they share insight, frameworks with each other*.
>
> *multidisciplinary: each member of the team operates within their own frame work and have their own goals. they refer the patient to each other without much discussion of the patient’s case”* (Female student, 2022-23).

Discordant rhetoric was identified in 15.9% of responses (20.0% in 2021–22, 13.7% in 2022–23, and 13.2% in 2023–24). These responses were not aligned with the definitions of one or both concepts found in the literature. Specifically, 9 (students did not provide a correct definition of interdisciplinarity, 11 did not correctly define multidisciplinarity, and 6 demonstrated an inaccurate understanding of both concepts. Examples of these discordant responses are presented below.

> *“Interdisciplinary means reaching out to others to come up with the best course of treatment for your patient. Multidiscipliniary means collaborating with people from different fields as they may have something to teach you that you have not included initially and it may help in a more comprehensive treatment and better prognosis overall for your patient”* (Male student, 2022-23).
>
> *“Interdisciplinary – involves various disciplines working on their own first, and then referring to one another. Multidisciplinary – involves multiple different disciplines (e*.*g*., *dentist, physiotherapist, nurse, medical doctor). Both are important!”* (Female student, 2021-22).

Switched ideas occurred in 18.3% of the responses (15.0% in 2021-22, 25.5% in 2022-23 and 15.1% in 2023-24), where students seemed to have swapped the definition of the two concepts. For example:

> *“Multidisciplinary care is more of a collaborative approach to care for a patient, whereas interdisciplinary care is care for a patient being provided by multiple disciplines independent of one another*.*”* (Male student, 2023-24).
>
> “*Interdiscipinary looks at each discipline individually and simply links the disciplinaries together, with no integration. Multidisciplinary involves the combination of multiple disciplines and allows each discipline to come together and share their perspectives on their field of expertise, utilizing each discipline fully*” (Female student, 2022-23).

Altered discourse observed in 20.1% of responses (28.3% in 2021–22, 11.8% in 2022– 23, and 18.9 in 2023–24). In these cases, the misunderstanding was not primarily related to the definitions of interdisciplinarity and multidisciplinarity themselves, but rather to the concept of “discipline.” Students appeared to equate disciplines with dental specialties rather than with different professions which is a key element in IPE. Examples of these altered understandings are presented below.

> *“Interdisciplinary is the work within a group of individuals who are all from the same field such as a group practice of general dentists. Multidisciplinary is the work amongst several dental specialties such as endodontics, prosthodontists, and so on*” (Male student, 2023-24).
>
> *“Interdisciplinary is when different dental specialties within the same field address the needs of the patient, for instance when a general dentist, periodontist and prosthodontist come together for treatment planning. Multidiciplinary is when these same specialists come together to create a wholesome and comprehensive plan addressing different body systems”* (Female student, 2021-22).

Inferential statistical analysis revealed that classes of 2023-24 and 2022-23 got more responses reflecting concordant knowledge while class of 2022-23 got more responses reflecting switched ideas; however, this difference was not statical significant (Table 1). Also, no significant differences were observed among the classes regarding age (p = 0.071), gender (p = 0.738), or theme (p = 0.189). In the logistic regression analysis, only age was associated with a higher probability of providing the correct definition as concordant knowledge. For each one-year increase in age, the odds of answering correctly increased by approximately 39% (adjusted OR = 1.39; 95%CI: 1.06–1.83; p = 0.017). However, although the result was statistically significant, the mean age varied very little between the groups, suggesting a very limited implication of this increase.

## Discussion

This study explored the extent to which senior dental students retained knowledge of multidisciplinarity and interdisciplinarity in the provision of collaborative, interprofessional care. Less than half of the students (45.7%) appeared to demonstrate an understanding of these concepts after learning about them. And approximately one in five students (20.1%) seemed to equate a discipline as a dental specialty, believing that interprofessional care occurs when different dental specialties collaborate. However, interdisciplinary collaboration among disciplines and the parallel practice of specialties within the same profession has been recognized as distinct in health profession education. Our finding suggests that some students may have difficulty differentiating collaboration across professions from collaboration among specialties within dentistry.[3,25]

Moreover, the extent to which the students are actually applying this knowledge in practice remains unknown. Knowledge alone does not consistently translate into behavioural change or improved interprofessional and team-based practices.[26] As a clinically focused profession, dentistry is delivered largely within silos, and collaborative, interdisciplinary practice is heavily influenced by systemic barriers, time constraints, and environmental factors. Such limitations restrict the implementation of meaningful changes required to support collaborative systems, even when there is an intention to do so.[27] Also, attitudes and behaviours are shaped by how knowledge is applied in practice, underscoring the importance of providing opportunities for collaborative clinical experiences beyond the traditional siloed approach to oral health care.[28,29] Furthermore, general dentists in Canada are expected to collaborate as members of healthcare teams and to ‘engage patients and others [health care providers] in developing plans that reflect the patient’s …. needs’.[30]

While IPE and awareness are foundational, long-term behavioural change requires attention to environmental contexts, self-regulation, motivation, and positive attitudes, together with opportunities for team-based care. Within the context of UBC’s health programs, IPE seeks to shift education away from a multidisciplinary approach, where physicians, nurses, dentists, pharmacists, and other professionals may independently review a patient’s case, toward an interdisciplinary model of care. Students are encouraged to integrate their expertise, actively involve patients, and participate in shared decision-making to improve overall health outcomes. In reality, however, dental students have relatively few opportunities to experience collaborative practice. This limited exposure may help explain why 54.9% of respondents either could not accurately define these terms, had forgotten their meanings, or may have never fully learned them in the first place, at least at a theoretical level.

Do these findings suggest that the substantial efforts undertaken by the university and faculty to integrate IPE into the undergraduate curriculum have been ineffective? The evidence suggests otherwise. Research demonstrates the value of preparing future healthcare professionals with the communication, leadership, and problem-solving skills necessary for effective teamwork central to IPE curricula.[3,6] However, these competencies must be applied through meaningful opportunities to work collaboratively with patients, families, and healthcare professionals from other disciplines in delivering coordinated and comprehensive care. Together, these educational approaches are designed to break down the silos that have traditionally separated oral healthcare, fostering a culture of mutual respect and shared values throughout professional training. They also help address challenges associated with chronic disease management and health inequities by promoting team-based models of care.[5] Nevertheless, significant challenges remain within the practice of dentistry.

At the University of British Columbia, IPE represents an effort to integrate interprofessional competencies throughout educational programs and practice environments during the early years of dental education. It also aims to help students recognize the benefits of collaborative care and respond to the evolving needs of increasingly diverse populations.[31] However, additional efforts are needed to provide team-based clinical opportunities that allow students to translate this knowledge into practice within interprofessional healthcare settings. Although dental hygiene students have participated in interdisciplinary care conferences focused on the health of long-term care residents,[26] opportunities for dental students to develop and apply interdisciplinary competencies remain few and far between, as we have recently discussed.[8] Collaborative care is known to help healthcare teams prioritize patient-centered care plans and improve the coordination of services.[32,33] Although IPE is primarily focused on undergraduate education, teaching and training institutions should also consider offering continuing education opportunities in interprofessional practice. Such opportunities could enable provide practicing clinicians to apply collaborative approaches in settings such as dental geriatrics [20,26,30] and hospital-based dentistry.[34,35] These efforts should complement, rather than replace, undergraduate IPE initiatives.

Despite the findings presented herein, this study has several limitations. Although it included three cohorts of students, it was conducted within a single dental program. As such, comparisons with other institutions are not warranted, and the generalization is limited. Information regarding student academic performance and assessment outcomes within the IPE curriculum itself was unavailable. Consequently, it was not possible to determine whether the student responses reflected learning gains, retention, loss of knowledge, or changes over time. That is, the data cannot distinguish between forgetting, misunderstanding, guessing, or never having learned the concepts. Because the question was embedded within an assessment, some students may have chosen not to answer the question, or run out of time. Nevertheless, all responses were included in the analysis to provide a comprehensive representation of student performance. Furthermore, because the question assessed knowledge of concepts rather than actual behaviours, the extent to which students can effectively collaborate in interprofessional settings remains unknown. Social desirability bias may also have influenced responses, as students may have provided answers that they believed instructors expected rather than expressing their genuine understanding. Future research should explore the extent to which dental students engage in interdisciplinary collaboration when given opportunities to do so. Follow-up studies should also investigate whether graduates apply the interprofessional knowledge and skills acquired once they enter clinical practice.

## Conclusions

Knowledge of interdisciplinary and multidisciplinary approaches appeared to be retained by some students. However, dental education programs, alongside other health professional training programs, should incorporate interprofessional education through both didactic and experiential learning opportunities to equip students with the skills necessary to provide collaborative care in future practice. A call to action is warranted for teaching and training institutions to consider offering continuing education opportunities in interprofessional education and collaborative practice throughout clinicians’ careers. Such initiatives, however, should complement rather than replace undergraduate IPE.

## Data Availability

All data produced in the present study are available upon reasonable request to the authors.

## Acknowledgements

We are grateful to the students whose answers to the question posed made this study possible. The authors would like to acknowledge the importance of the IPE curricula that students experience in their junior years, supporting their student learning. The authors declare no conflicts of interest and confirm accountability for all aspects of the work.

